# Association of acute kidney injury with the risk of dementia: A meta-analysis protocol

**DOI:** 10.1101/2021.07.27.21261192

**Authors:** Salman Hussain, Ambrish Singh, Benny Antony, Jitka Klugarová, Radim Líčeník, Miloslav Klugar

**Author notes:** Corresponding author Dr. Miloslav Klugar, Czech National Centre for Evidence-Based Healthcare and Knowledge Translation (Cochrane Czech Republic, Czech EBHC: JBI Centre of Excellence, Masaryk University GRADE Centre), Institute of Biostatistics and Analyses, Faculty of Medicine, Masaryk University, Brno, Czech Republic.

## Abstract

Acute kidney injury (AKI) is a complex disorder characterized by an abrupt decline in kidney function over a short period of time. Published epidemiological studies linked AKI with the development of dementia. This meta-analysis aims to understand the pooled risk of dementia in AKI patients compared to non-AKI patients. MEDLINE and Embase databases, and the grey literature in five sources were searched to identify the studies assessing the association of AKI with dementia. The Newcastle-Ottawa scale (NOS) will be used to determine the quality of included studies. The primary outcome of this study will be the risk of dementia among AKI patients compared to non-AKI patients. Subgroup analysis and sensitivity analysis will also be performed. Review Manager version 5.4.1 will be used to perform the meta-analysis.

## Background

Acute kidney injury (AKI) is a complex disorder characterized by an abrupt decline in kidney function over a short period of time. [1] AKI is associated with poor quality of life, decreased productivity, and negative health economic impact. [2,3] Published epidemiological studies linked AKI with the development of dementia. [4,5] Dementia is a neurodegenerative disorder characterized by progressive deterioration of intellectual function, and its prevalence raised considerably in the last few decades. [6] The occurrence of dementia in patients with AKI is of clinical importance as dementia is associated with an increased humanistic and economic burden. [6]

The preliminary search of existing systematic reviews was performed on July 2021 in Epistemonikos, PROSPERO, Open Science Framework, Cochrane Library and JBI Evidence Synthesis and no reviews evaluating the association of acute kidney injury with the risk of dementia were identified. Hence, it is of importance to understand the pooled risk of dementia in patients with AKI.

## Objective

This meta-analysis aims to assess the association between AKI and the risk of dementia.

## Methods

This meta-analysis will be conducted and reported according to the recommendations of the preferred systematic review and meta-analysis (PRISMA) [7] and Meta-analysis of Observational Studies in Epidemiology (MOOSE) reporting guidelines. [8]

## Search strategy

A three-step search strategy was already utilized to locate both published and unpublished studies for this systematic review and meta-analysis. An initial limited search was undertaken in MEDLINE (Ovid) using keywords and index terms related to AKI and dementia. An analysis of the text words in the title and abstract and the index terms used to describe the articles were followed. A second search using all identified keywords and index terms was conducted in MEDLINE (Ovid) and Embase (Ovid) databases (the search period was from inception to 14^th^July 2021). Thirdly, the reference lists of all studies that met the inclusion criteria will be checked for additional records. Lastly, abstract booklets of major international nephrology and neurology congress: World Congress of Nephrology, American Society of Nephrology, European Renal Association – European Dialysis and Transplant Association (ERA-EDTA), Asian Pacific Congress of Nephrology, Neuroscience, Alzheimer’s Association International Conference (AAIC), and American Academy of Neurology from last two years will be searched. We will also do the citation tracking of all the qualified articles for inclusion.

## Study selection/Eligibility criteria

### Characteristics of participants (P)

We will include patients with AKI with a confirmed diagnosis of dementia. Primary studies including individuals with AKI with dementia at the entry of the cohort were excluded from the analysis

### Characteristics of exposures (E)

This study will focus on patients exposed to AKI.

### Outcomes of interest (O)

Our primary outcome is to assess the risk of dementia among patients with AKI.

### Study design/type of studies

Observational analytical studies (prospective, retrospective, cohort, or case-control) that assessed the risk of dementia in the AKI population compared to the risk of dementia in the population without AKI will be included.

All the retrieved articles will be assessed for inclusion based on title and abstract screening, followed by full-text screening by two independent reviewers. The discrepancy in study inclusion will be resolved by discussion with the third reviewer.

## Quality assessment

The Newcastle-Ottawa scale (NOS) will be used to evaluate the quality of included studies. Each article will be assessed independently by two researchers using the NOS. A study can achieve a maximum of 4 points in the selection, 2 points in the comparability, 3 points in exposure (case-control studies), or outcome (cohort studies) domain of the scale. Studies with 8-9 scores, 7-8 scores, and ≤6 scores will be classified as high, medium, and low-quality studies. All studies will be included in the meta-analysis regardless of their methodological quality, however the influence of methodological quality on the outcome will be explored by the sensitivity analyses.

## Data extraction

All the relevant data related to the population, intervention, comparator, and outcomes will be captured. Both the adjusted and unadjusted outcomes will be extracted if reported in the included studies. Data extraction will be independently performed by two reviewers.

## Statistical analysis

Review Manager version 5.4.1 will be used to perform the meta-analysis. A generic inverse variance method will be applied to compute the overall risk ratio (RR) of dementia in AKI patients. As the event is rare, the RR, odds ratio, and hazard ratio will be considered as an equivalent measure of risk; therefore, we will use RR representing all these measures for simplicity. [9] Heterogeneity will be assessed using I^2^ value Cochrane chi-square value. Cochrane chi-squared value (p< 0.10) and I2 value ≥50% may represent considerable heterogeneity. [10] Considering the high probability of between-study variance due to distinction in populations and techniques used to diagnose AKI and dementia, the random-effect model will be chosen over the fixed-effect model. Sensitivity analysis will be performed by omitting one study at a time to understand the impact of any single study on the effect size. Similarly, subgroup analysis will also be performed. The potential for publication bias will be assessed using a funnel plot construction if there are ten or more studies. The y-axis will be the study population size, and the x-axis will be the measure of effect for that property.

## Certainty of evidence

As per COSMIN guidelines for systematic reviews, the quality of evidence will be assessed with the Grading of Recommendations Assessment, Development and Evaluation (GRADE) using the five domains for decreasing certainty of evidence: risk of bias, consistency, directness, precision, and publication bias, and three domains for increasing certainty of evidence: large effect, dose-response gradient and plausible confounding factors.[11] Reviewers assessing the certainty of evidence will refer to the GRADE handbook [12] to ensure proper utilization of the tool for systematic reviews of exposures. The overall certainty of evidence will be considered high (a lot of confidence that the true effect is similar to the estimated effect), moderate (the true effect is probably close to the estimated effect), low (true effect might be markedly different from the estimated effect), or very low (the true effect is probably markedly different from the estimated effect).

## Data Availability

The data that support the findings of this study will be available from the corresponding author upon a reasonable request.

